## Supplementary figures and images for "Herpesvirus Reactivations in Critically-Ill COVID-19 Patients with Autoantibodies Neutralizing Type I Interferons"

### Supplemental Figure 1

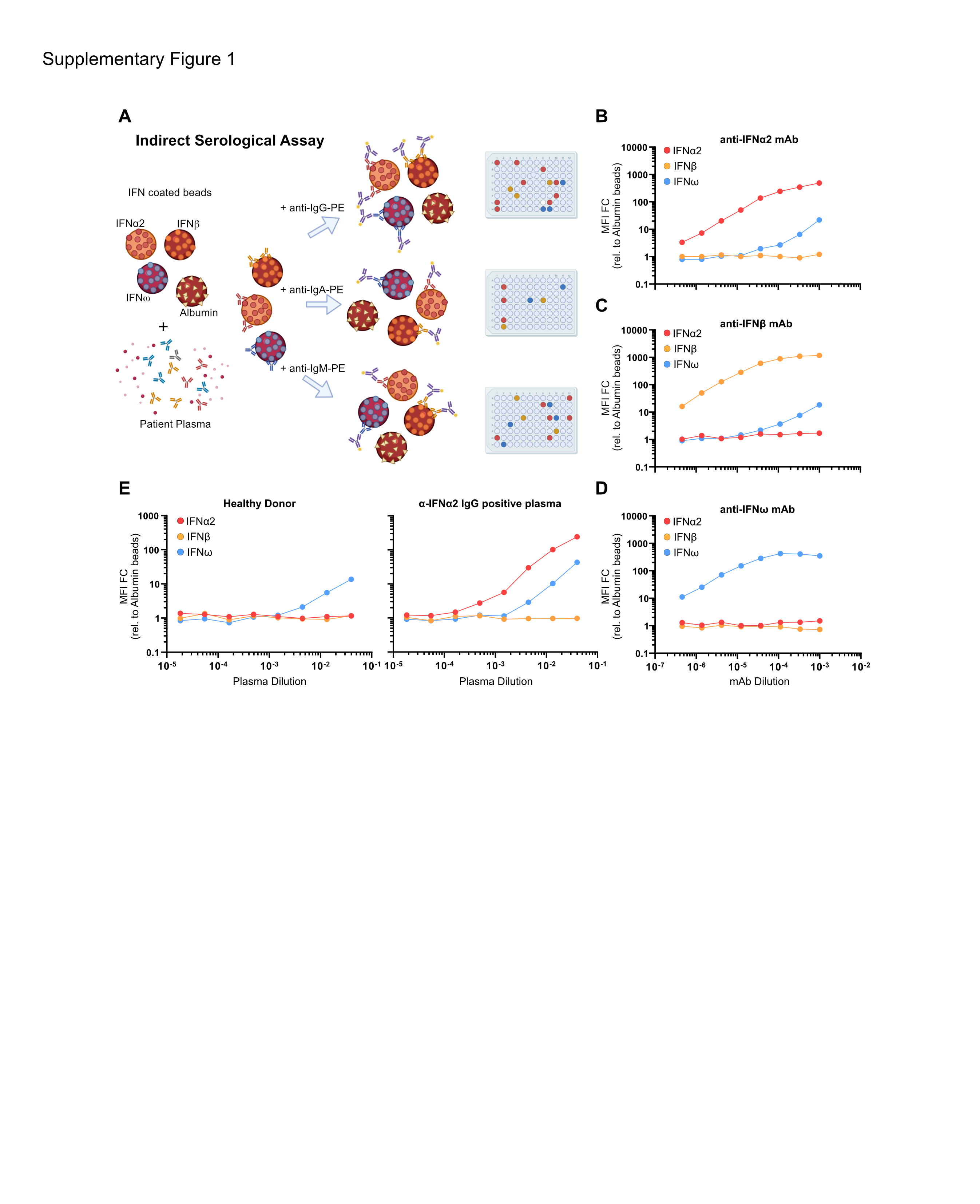

### Supplemental Figure 2

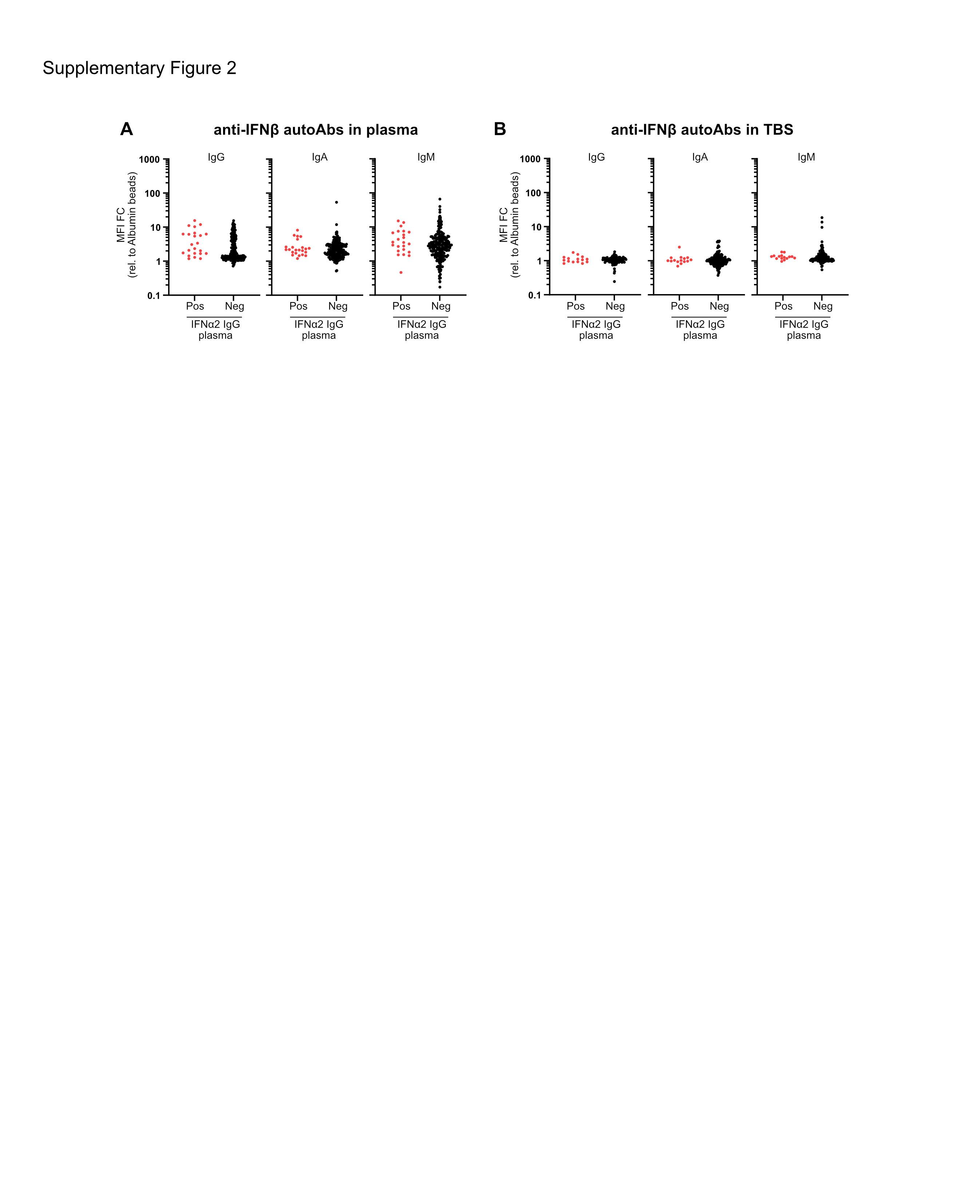
